# Exploring the use of social media and online methods to engage knowledge users in creating research agendas: Lessons from a pediatric cancer research priority-setting partnership

**DOI:** 10.1101/2022.12.12.22283382

**Authors:** Kyobin Hwang, Surabhi Sivaratnam, Rita Azeredo, Elham Hashemi, Lindsay A. Jibb

## Abstract

While social media is increasingly leveraged to engage knowledge users in research priority-setting, there remains sparse explicit descriptions on how to implement it to build knowledge user-led research agendas. The aim of this study was to review a case study where social media was utilized to engage Canadians within the pediatric cancer community in a research priority-setting exercise; specifically highlighting the social media-based recruitment process, including recommendations on how to optimally engage key potential participants. A priority-setting partnership was launched to develop a stakeholder-driven research agenda in pediatric cancer in Canada. Social –media-based strategies were implemented for participant recruitment, developing a website, launching graphics and advertisements, and engaging internal and external stakeholders. These strategies incorporated the use of various social media platforms. We used descriptive statistics to analyze the data, in addition to the analytics provided by the platforms mentioned. Throughout the duration of the PSP, we garnered 870 Instagram followers, 450 Twitter followers, 69 Facebook page likes, 27 TikTok followers, 20 LinkedIn followers and 789 unique visitors to our website. Our Facebook page reached 28,641 people, while our Instagram profile reached 2,954 people. This social media campaign resulted in 330 individuals completing the initial survey of the PSP, and 197 individuals completing the interim prioritization survey. Social media is a novel approach to engage stakeholders in the development of a research agenda. Our study identified the following strategies as effective in increasing participation in our PSP: (1) creating a unified brand, (2) prioritizing accessibility (e.g., providing alternative text for all images), (3) ensuring social media campaign is reflective of the target audience by diversifying platforms and intermittently tailoring content to specific populations, (4) optimizing campaign’s reach via paid advertisements and circulating promotional material to partner organizations and individuals for them to subsequently share with their networks.

**AUTHOR SUMMARY:** Our study evaluated the usage of social media to engage pediatric cancer patients, survivors, their family caregivers, and healthcare providers in setting research priorities for the field of pediatric oncology. As a resource for other research teams, we offer a description of the traction gained by our social media campaign and factors contributing to an increased engagement in the priority-setting process. Social media-based strategies were utilized for participant recruitment; this included developing a website, launching graphics and advertisements, and engaging various stakeholders. Throughout the study duration, we gained 870 Instagram followers, 450 Twitter followers, 69 Facebook page likes, 27 TikTok followers, 20 LinkedIn followers, and 789 unique visitors to the study website. Social media is a relatively new approach to engage individuals for research priority-setting. Our study identified the following strategies as effective methods to increase engagement: (1) creating a unified branding, (2) prioritizing accessibility (e.g., providing alternative text for all images), (3) using various platforms and tailoring content to specific populations, and (4) optimizing campaign reach through paid advertisements and by circulating promotional material to partner organizations and individuals to share within their networks. Further investigation of the privacy implications of social media use for priority-setting research is needed.

## BACKGROUND

In recent years social media has gained an important role in healthcare, including engaging key healthcare knowledge users in research (1,2). Researchers are increasingly utilizing Facebook, Twitter, and YouTube to enable participant recruitment into research studies (3,4,5) and to disseminate research findings (6). Social media-based methods have also been increasingly used to enable the engagement of individuals with lived healthcare experiences (i.e., patients, family members, clinicians, and other advocates) in setting research priority agendas (7). Historically, such agendas have been set with little reference to, or input from, those with lived experience—limiting the capacity to effectively translate research findings into practice and policy (8,9.10). Consequently, there is a risk that without the active engagement of patients, families, healthcare professionals and other advocates in research priority determination, researchers may face an unfortunate cycle of producing underutilized evidence, with a resulting loss of public and professional trust and limited impact on patient outcomes.

In light of the critical need to engage those with lived experience in research agenda building and the potential opportunities to support such work through online methods, our research team utilized social media, amongst other online tools, as a key method enabling knowledge user engagement in our pan-Canadian pediatric cancer research priority setting partnership (PSP). In brief, through our PSP we surveyed the Canadian childhood cancer community to elicit their research questions, and subsequently engaged the group in a priority setting activity to identify an inclusive childhood cancer research agenda. To create such an inclusive agenda, we aimed to engage a diverse group of child patients, pediatric cancer survivors, as well as their family members and healthcare professionals. This recruitment occurred within the bilingual (English and French).

While social media is more routinely leveraged to engage knowledge users in research priority-setting, there remains sparse explicit descriptions of the methodology and how best to implement it to effectively build knowledge user-led research agendas (7). Here we review a specific case study where social media and other online modalities were leveraged to engage Canadians within the pediatric cancer community in a research priority-setting exercise. As a resource for other research teams, we offer a description of our social media-based recruitment process, engagement with our social media campaign, and factors associated with increased engagement in our priority-setting process. We also discuss the limitations of our efforts and make recommendations on the specifics of social media use to engage key knowledge users in PSP.

## METHODS

### Overview of Pediatric Priority Setting Partnership

We followed the James Lind Alliance process for the development of stakeholder-engaged research agendas (11). At the onset of our PSP, a steering group, comprised of childhood cancer survivors, family members, and pediatric oncology clinicians was established. This steering group guided all operational decisions of our project. The first step of our approach involved launching a national bilingual (French and English) online survey in the Winter of 2020 to collect research questions from the Canadian pediatric cancer community. Collected questions were collapsed and collated into summary questions, which were checked against the extant scientific literature. Adequately addressed questions were removed from the question roster and a second national, bilingual online survey was launched in the Winter of 2021 requesting children with cancer, survivors, family members and health professionals to prioritize the unanswered research questions. A shortlist of these questions was then taken to a pan-Canadian consensus-building workshop conducted in March 2022, where the top ten research priorities in Canadian pediatric oncology were established. A full description of methods and results for this PSP is reported elsewhere. Social media and other online tools were utilized to promote both of our online surveys.

### Preparatory Activities for Social Medial Campaign

#### Scoping review

Prior to launching our social media campaign, a scoping review was conducted to describe (1) the existing literature on social media–based strategies used to enhance knowledge user participation in health research priority setting, (2) the recommendations for social media–based research priority-setting campaigns, (3) the benefits and limitations of the method, and (4) the recommendations for future campaigns (7). We identified a total of 23 papers reporting on 22 unique studies. Our central findings included identification of the social media platforms and online tools used to engage with health research stakeholders, the related online engagement strategies utilized, methods to assess social media campaign effectiveness, and novel suggestions to enhance engagement in priority-setting. The results of this review informed the social media strategies implemented in this study.

#### Creation of social media content

An initial branding package was created in Adobe Photoshop (CC 2017; Adobe, San Jose, CA). Two trained graphic designers created this package using brand-unifying fonts, colour palette, and vector graphics. During the development of these graphics, the designers utilized WebAim (https://webaim.org/resources/contrastchecker/) to ensure all graphics fulfilled the minimum colour contrast requirements for web and mobile accessibility. This package was then reviewed by the PSP steering group to ensure the content and design was appropriate for the targeted audience of patient-, survivor-, family-, and clinician-knowledge users. Steering group suggestions were used to modify and finalize the branding package. This branding scheme was then used as the basis of a hub of static graphics and videos that would be utilized in the social media campaign. The corresponding text for social media posts, calling for engagement in the PSP, was created by a researcher trained in marketing, in both English and French.

#### Establishing analytic tools

Dedicated social media accounts were created on Facebook (www.facebook.com), Twitter (www.twitter.com), Instagram (www.instagram.com), LinkedIn (www.linkedin.com), YouTube (www.youtube.com), and TikTok (www.tiktok.com), ensuring that whenever applicable, accounts were registered under the “business/professional” account types. This registration type enabled the option of viewing social media analytics, which included overviews of the demographic characteristics of those engaging with our social media content (i.e., age, gender, education levels, job titles, location, language) and page insights (i.e., likes, comments, shares, page views, page traffic and activities-including the length of time of individual sessions). We also created a Hootsuite account (Hootsuite Inc), which provides access to aggregated analytics of all user profiles within various networks, as well as additional analytics (i.e., most common times of day during which followers interact with social media accounts/pages). The Hootsuite platform also enabled the research team to schedule posts, and provided a single point to launch posts across various social media platforms.

### Social Media Engagement

#### Initial launch of social media campaign

A series of introduction posts were generated with the intention of establishing credibility related to our campaign. The first post identified the research project members, including the research team and the steering group, and subsequent posts introduced the concept of a PSP. At the initial launch of our social media accounts, we contacted key childhood cancer organizations and social media users through direct messaging to introduce the PSP. We further began following these users in an effort to increase the number of users interacting with the established social media accounts.

#### Social media campaign

The content of the social media posts included reviews of the goals of the PSPs, the key personnel involved, calls for action (i.e., encouraging users to complete each survey), and, intermittently, posts relevant to the pediatric oncology community (e.g., recognizing childhood cancer awareness month). Posts typically included graphics in the form of photos, videos, and graphics interchange format (GIFs), and associated text included hashtags, links (i.e., to the project website or the survey), tags, and various relevant emojis. Sample graphics are shown in Figure 1a and 1b. Throughout the social media campaign, we monitored (1) social media analytics to identify gaps in interaction with campaign and (2) demographics self-reported by survey participants to identify gaps in communities of individuals completing each of the surveys. The identification of these gaps subsequently informed the content of future social media posts. For example, during the campaign, these metrics helped the research team identify a decreased response rate from certain populations, including Black, Indigenous, People of Colour (BIPOC), French-speaking individuals, and male-respondents, and efforts were made to target graphics towards these populations. We also monitored social media analytics, which provided optimal timing for posts (i.e., the times at which the majority of our audience was online and interacting with our posts). Posting schedule was continuously adjusted to reflect these optimal times. Sample targeted graphics are shown in Figure 2a and 2b.

**Figure 1a and 1b:**
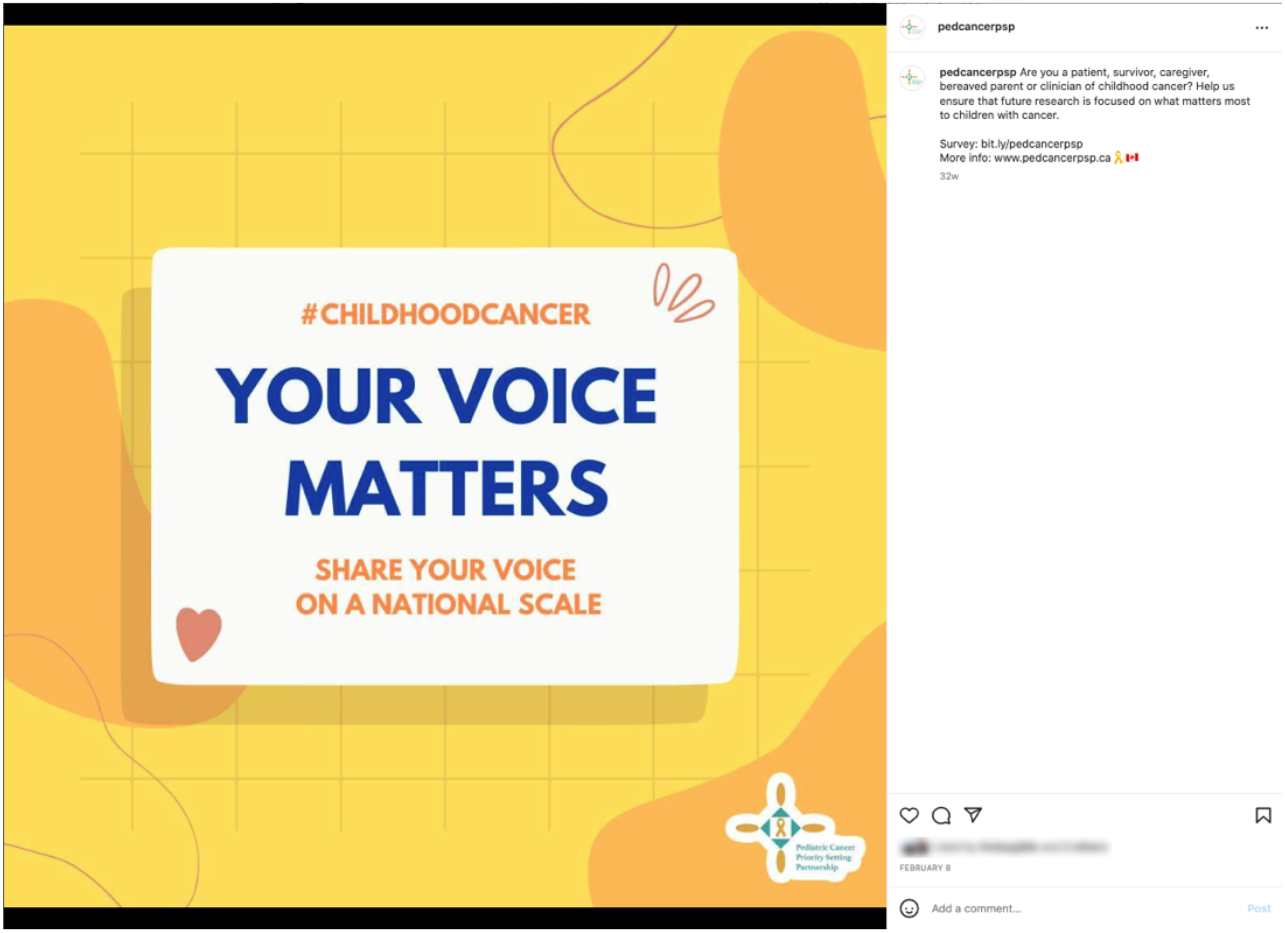
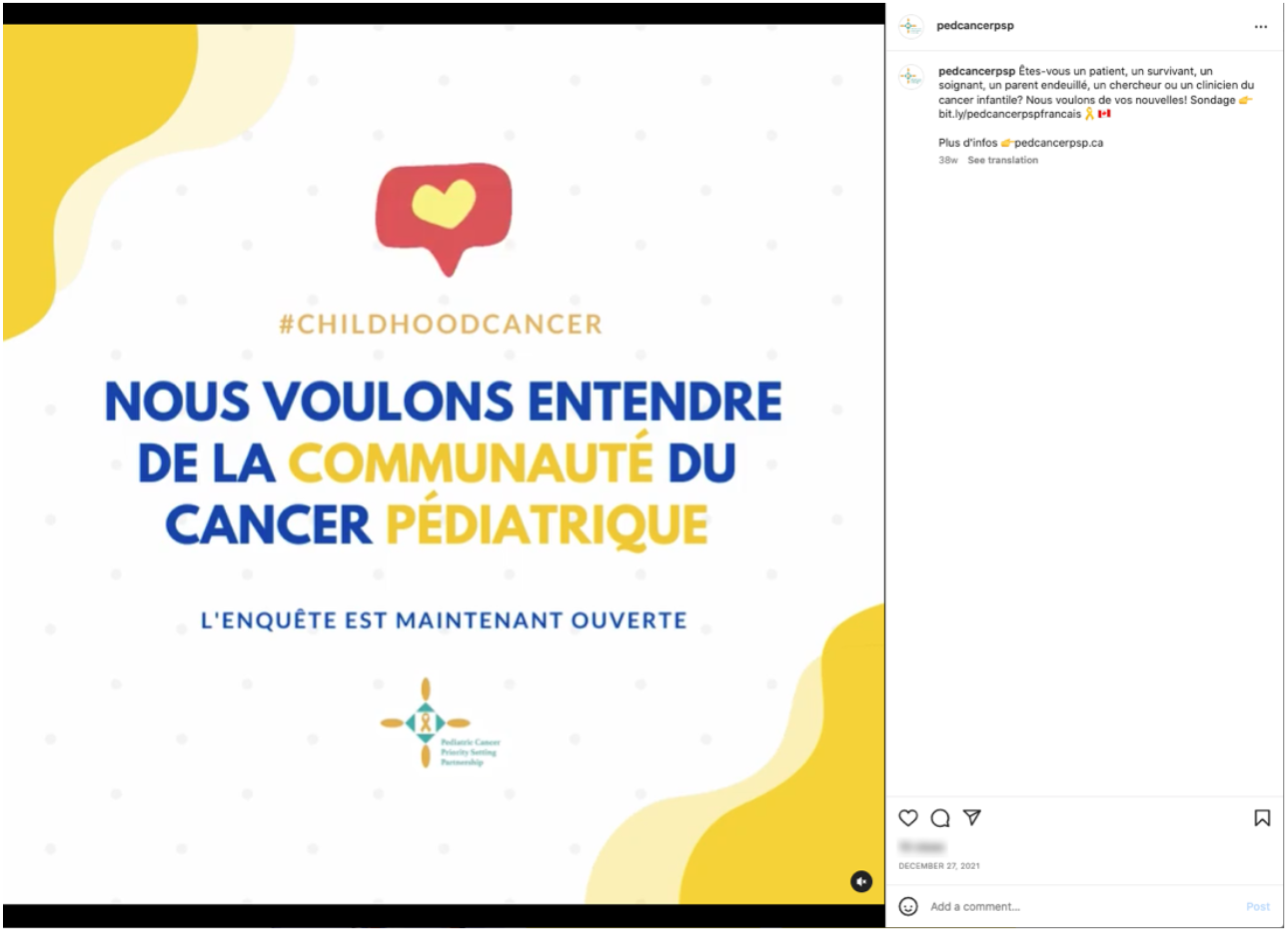
Example of graphics from social media campaign

**Figure 2a and 2b:**
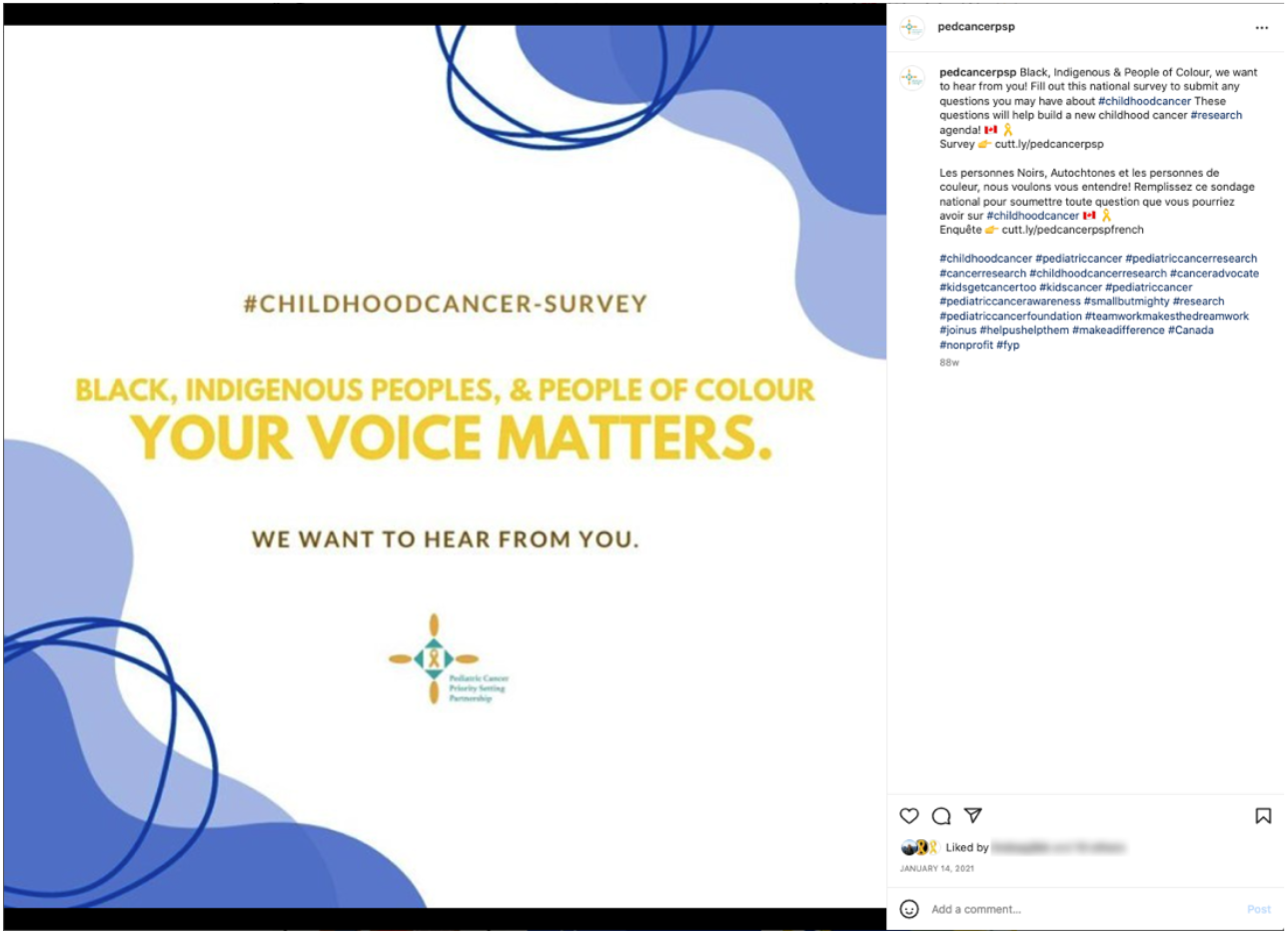
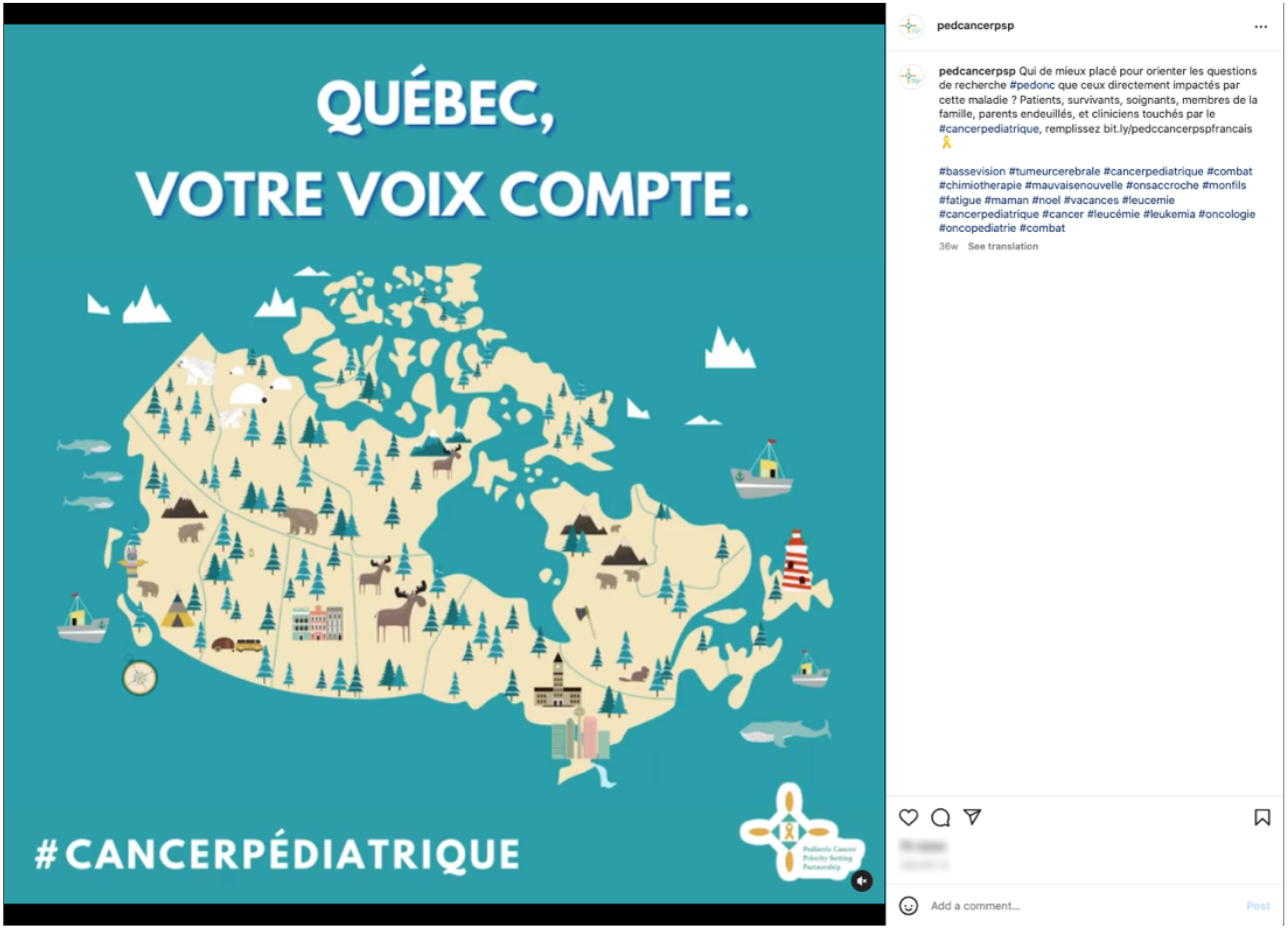
Example of graphics targeting specific groups

#### Paid advertisements

During the social media campaign, we periodically implemented paid advertisements on Facebook, Instagram and Twitter. Our team closely monitored advertisement associated analytics to identify ways to augment the campaign. For example, we identified that video posts were more cost-effective than image-based posts, and all future advertisements were then solely accompanied with videos. Each social media platform required specifications on ad delivery, ad content, design language, targeted audience, including location, and dates of deployment. We also targeted paid advertisements towards the aforementioned underrepresented groups, by augmenting the “target population” options within the advertisement settings (e.g., to target French-speaking individuals, we restricted advertisements to be shown to Quebec residents only).

### Additional Online Engagement Strategies

#### Website

Prior to launching our social media accounts, a website for the research project was created (www.pedcancerpsp.ca). This website was created using Wix (Wix.com Ltd). During the development of the website, accessibility needs were optimized by ensuring alternative text was available for all graphics. The website was also developed within the desktop and mobile version to ensure all devices could access our website. Finally, utilizing Wix’s search engine optimization (SEO) checklist, the website was set up to ensure it could be found via search phrases on various search engines. The SEO checklist included the following components: (1) set the homepage’s title for search results, (2) add the homepage’s description for search results, (3) update the text on website’s homepage, (4) make website’s homepage visible in search results, (5) optimize website for mobile devices, (6) connect website to a custom domain, and (6) connect your site to a google search console. The website provided a centralized location for prospective participants to learn more about the project, the James Lind Alliance methodology, the research team, and registration to our mailing list.

#### Engagement via internal and external stakeholders

Throughout the social media campaign, we contacted external stakeholders, including Canadian childhood cancer organizations, advocacy groups with prominent social media accounts, and non-governmental organizations affiliated with pediatric cancer, and subsequently requested their assistance in promoting the project. Contact methods included email correspondence, as well as direct messaging via social media platforms. When such individuals and organizations agreed to help with survey promotion, we provided them with a template package, which included tailored graphics, pre-written messaging, sample newsletter graphics, and draft emails that they may send to their contact lists. A similar methodology was implemented when engaging with internal members (i.e., members of the research team and members of the steering committee).

### Data Analysis

Facebook, Instagram, and Twitter provide analytics to page administrators, allowing for monitoring and assessment of the usage of a page; a similar tool is available on Wix. These analytics allowed the research team to measure user engagement within each online platform. For Instagram, Facebook, and Twitter, we monitored reach, impressions, and clicks. For our website, we monitored unique visitors, site sessions, and page views. We also collected demographic information from our survey. Overall, we used descriptive statistics to analyze the data.

## RESULTS

### Initial research priority-setting survey

The initial research question gathering survey was available for completion from December 7, 2020, to March 3, 2021. A total of 330 individuals responded to our initial survey, of which 80 (23.9%) were childhood cancer patients and survivors, 179 (53.4%) were family members and friends of children who have or had cancer, and 76 were healthcare professionals (22.7%). The majority of family members/friends identified as mothers (n=161; 89.9%), followed by fathers (n=13; 7.3%), then friends (n=2; 1.1%). A range of multidisciplinary professionals responded, including registered nurses (n=32, 42.1%), oncologists (n=22, 28.9%), physical therapists (n=3, 3.9%), and psychologists (n=3, 3.9%). Majority of responded lived in Ontario (n=160, 47.8%), with the second largest group living in British Columbia (n=54;16.1%). There was representation from all of Canada’s provinces, though there were no response from territory residents.

### Interim prioritization survey

The interim research question prioritization survey was available for completion from January 10, 2022, to February 19, 2022. A total of 197 individuals responded to our interim prioritization survey, of which 49 (24.5%) were children or survivors of childhood cancer, 102 (51.0%) were family members or friends of a child that was diagnosed with cancer, and 49 (24.5%) were healthcare professionals who take care of children with cancer or survivors or do work in research related to pediatric cancer or survivorship. More nurses (n=24, 49.0%) responded than medical doctors (n=10, 20.4%); and there were 6 (12.2%) respondents that were research personnel and 6 (12.2%) selected ‘Other.’ The majority of survey respondents were once again Ontario residents (n=84, 42.0%), followed by Quebec (n=59, 29.5%) and Alberta (n=16, 8%). Similar to the initial survey, there was representation from all the provinces, though there were no respondents from the Northern territories.

### Social media campaign overview

Over the course of our entire PSP —from April 2020 to September 2022—we garnered 870 Instagram followers, 450 Twitter followers, 69 Facebook page likes, 27 TikTok followers, and 20 LinkedIn followers. Social media campaigning efforts were prioritized on Facebook, Twitter, Instagram, as well as our study website. Figures 3a and 3b provide an overview of how survey response rates correspond to key timepoints and efforts during the social media campaign period.

**Figure 3a and 3b.**
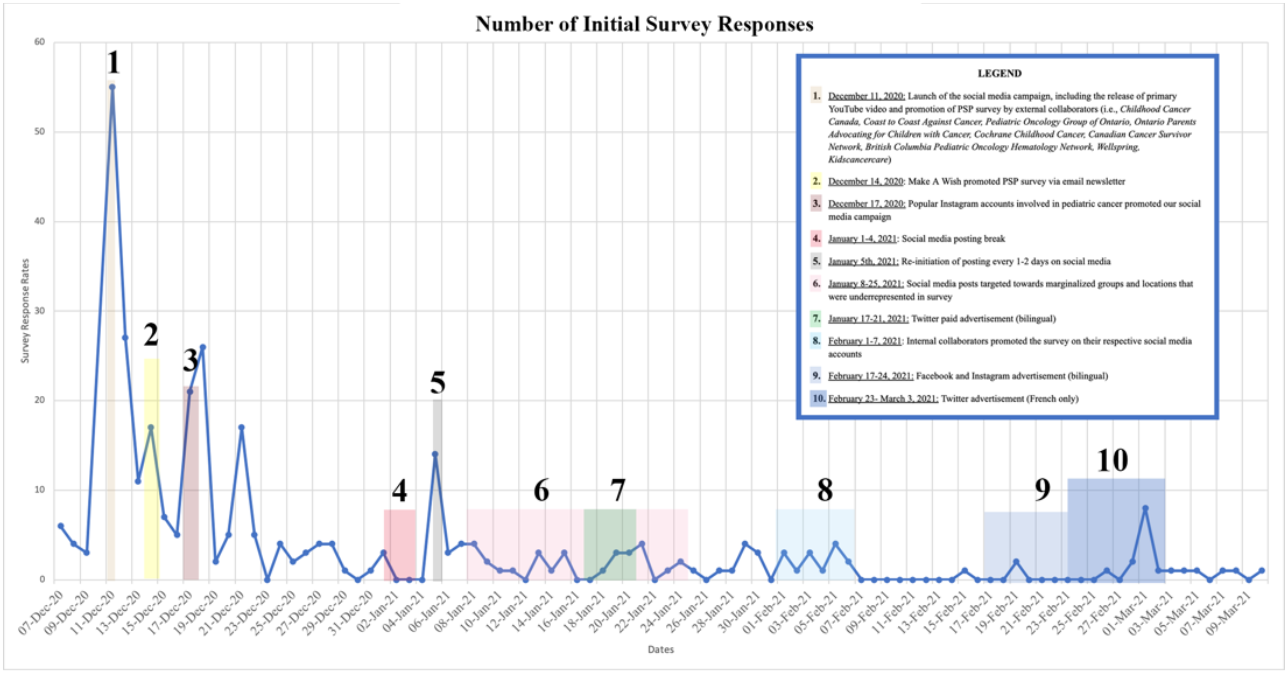
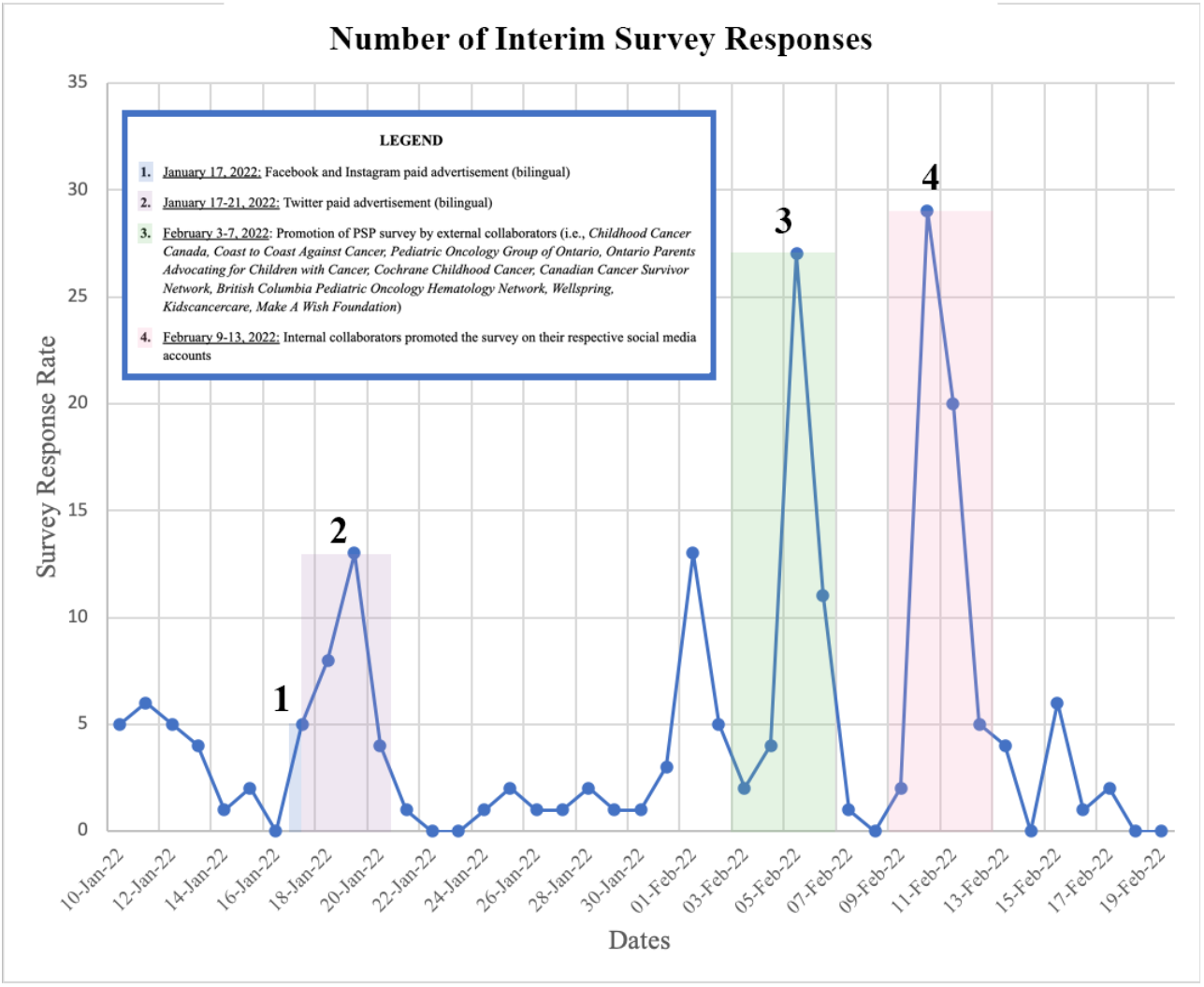
Survey response rates across social media campaign period

### Study website engagement

The study website received 1029 site sessions (visits ended after 30 minutes of inactivity), with 53% of users (n=547) accessing the site via desktop, 46% via a mobile device (n=470), and 1% via tablet (n=12). Figure 4 depicts the site session trend over time. Peak site sessions occurred between December 2020 to March 2021, as well as January 2022 to March 2022—time periods that corresponded to the interval during which each survey was open.

**Figure 4.**
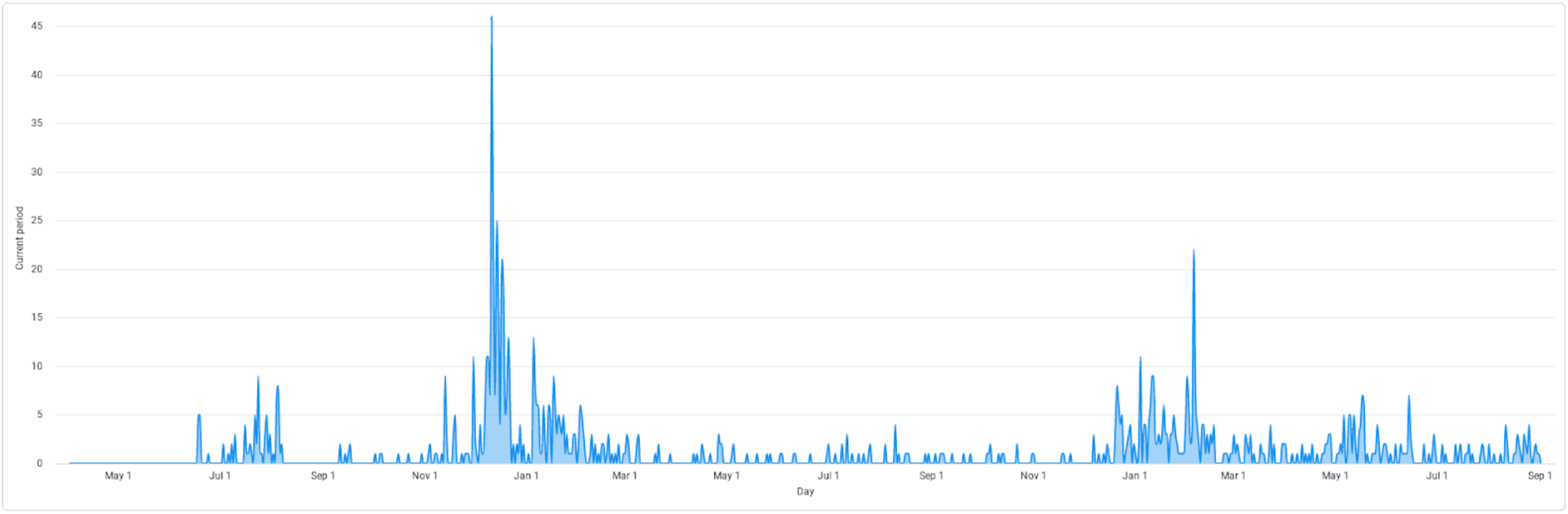
Site session trend over social media campaign period

The website garnered a total of 789 unique visitors, with most users accessing the website from Canada (n=622, 78.8%), followed by the United States (n=73, 9.3%). Figure 5 depicts a map with the traffic by location. The majority of unique visitors were from social media (n=148, 32.7%), followed by direct referrals (n= 146, 32.2%), organic searches (n= 105, 23.1%), and referrals (n= 54, 11.9%). Direct referrals are defined as visitors that typed our site address directly into their browser, without clicking any link. Organic searches are defined as visitors that clicked an organic search result from a search engine, such as google. Referrals are defined as visitors that clicked a link placed on another website that is not a search engine or a social media platform. Table 1 provides more information on the specifics of the traffic source.

**Table 1.** Overview of website traffic sources

| Traffic category | Traffic source | Site sessions | Page views | Unique visitors |
| --- | --- | --- | --- | --- |
| Direct | N/A | 186 | 203 | 146 |
| Organic search | Google | 161 | 199 | 94 |
| Social | Instagram | 59 | 66 | 51 |
| Social | Twitter | 56 | 66 | 42 |
| Social | Facebook | 47 | 58 | 46 |
| Referral | sickkids.ca | 26 | 26 | 25 |
| Referral | redcapexternal.research.sickkids.ca | 19 | 22 | 17 |
| Email | Email | 15 | 16 | 11 |
| Organic search | DuckDuckGo | 5 | 10 | 4 |
| Social | LinkedIn | 9 | 10 | 8 |
| Referral | Linktree | 5 | 5 | 5 |
| Organic search | Bing | 4 | 4 | 4 |
| Referral | pogon.convio.net | 2 | 3 | 2 |
| Referral | veeva.io | 2 | 2 | 2 |
| Organic search | Yahoo | 2 | 2 | 2 |
| Social | YouTube | 1 | 1 | 1 |
| Organic search | ecosia.org | 1 | 1 | 1 |
| Referral | outlook.live.com | 1 | 1 | 1 |
| Referral | us13.admin.mailchimp.com | 1 | 1 | 1 |
| Referral | com.linkedin.android | 1 | 1 | 1 |

**Figure 5.**
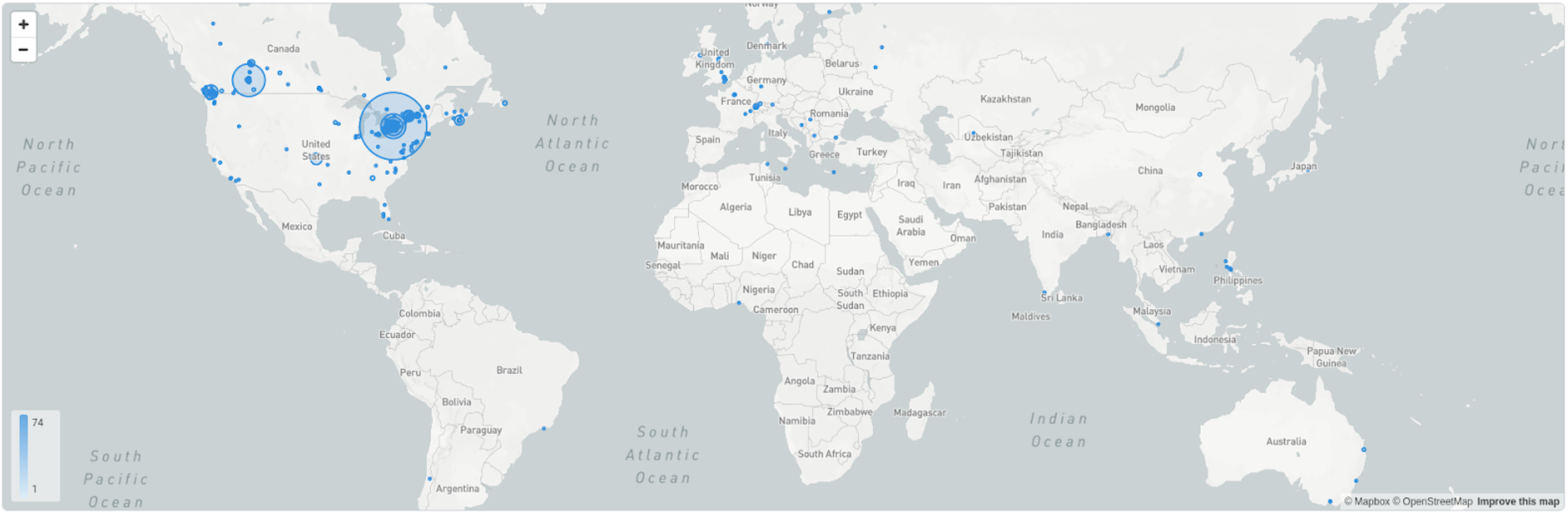
Geographic distribution of website users

### Facebook, Instagram and Twitter-focused campaign engagement

Over the course of the project, we made 152 posts on Facebook and 148 posts on Instagram. Our Facebook page was visited 426 times, while our Instagram profile had 1,893 visits. The reach of our Facebook page, or the number of individuals seeing any content from or about the page (including from others who interact with the page), was 28,641 people and our Instagram profile reach was 2,954 people, as seen in Figure 6. Considering activity related to our paid advertisements, over both Instagram and Facebook, our social media campaign’s paid content reach amounted to 24,137, while paid impressions (which may include multiple views of a post by the same individual) corresponded to 40,721. Paid reach compared to amount spent can be viewed in Figure 7.

**Figure 6.**
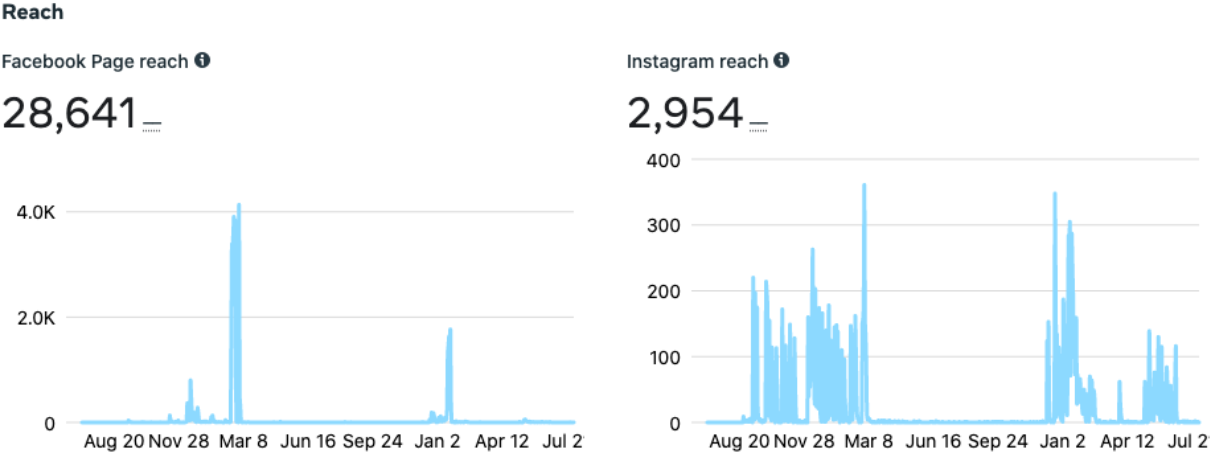
Facebook and Instagram reach

**Figure 7.**
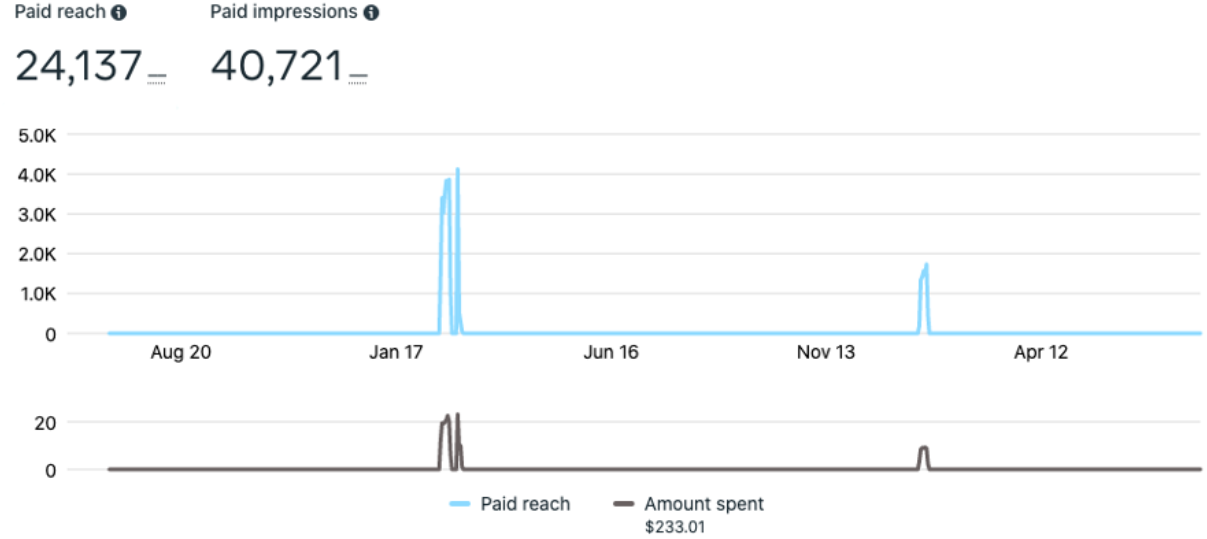
Paid advertisement trends

Our Twitter page garnered 452 followers. The greatest number of impressions, defined as the number of times an individual viewed a tweet, during a 90-day period occurred in January to March 2021—a time period within which both organic posts and paid advertisements were implemented. Table 2 provides the number of impressions during 90-day time periods, the number of daily impressions, and details of the trend of impressions over the time period.

**Table 2.**
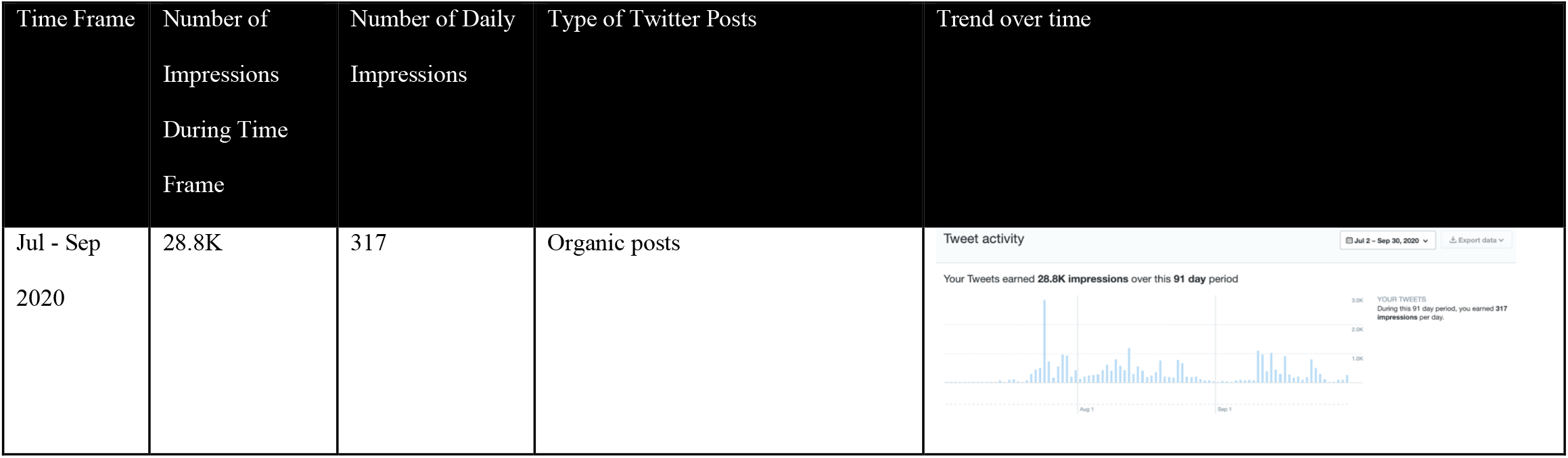
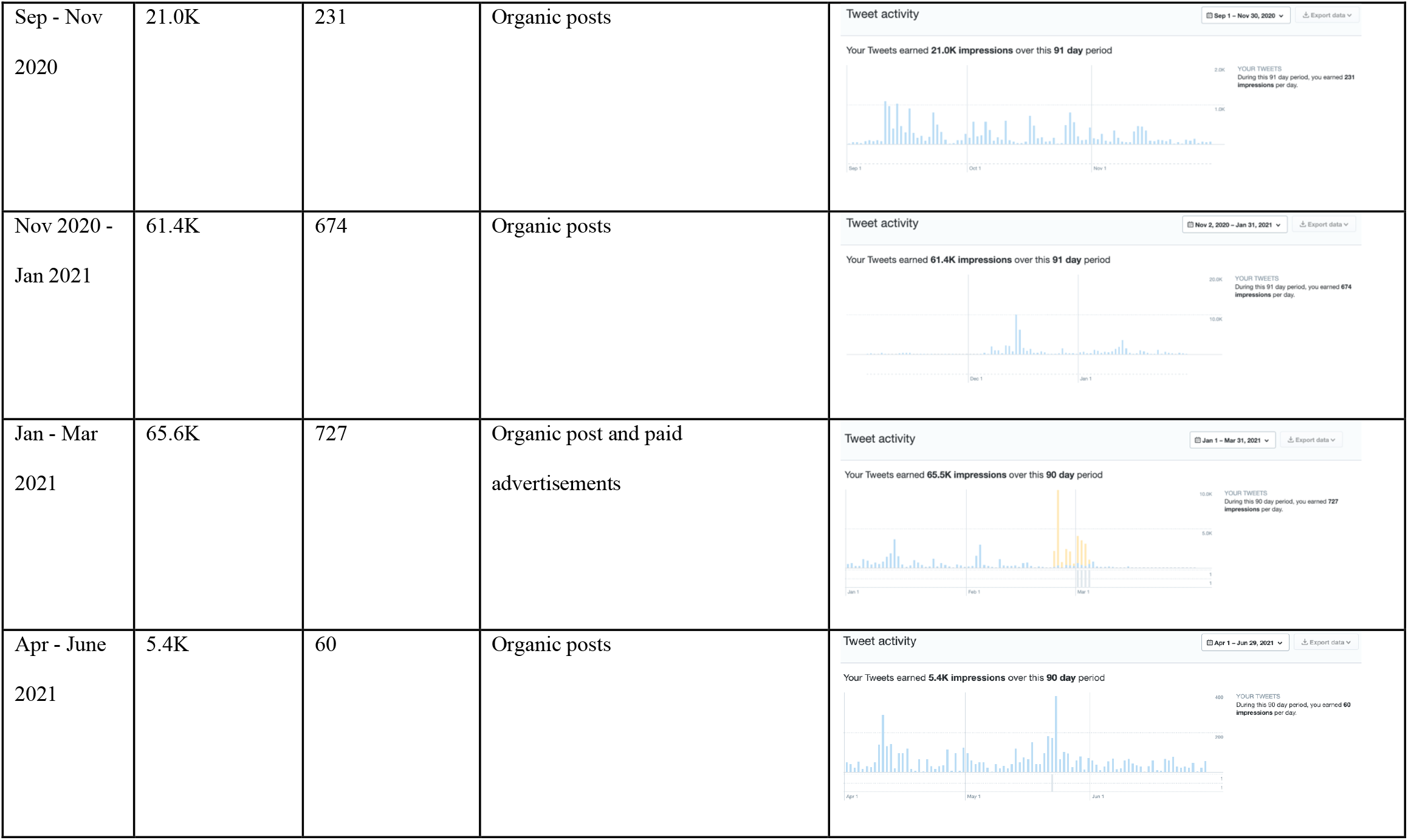
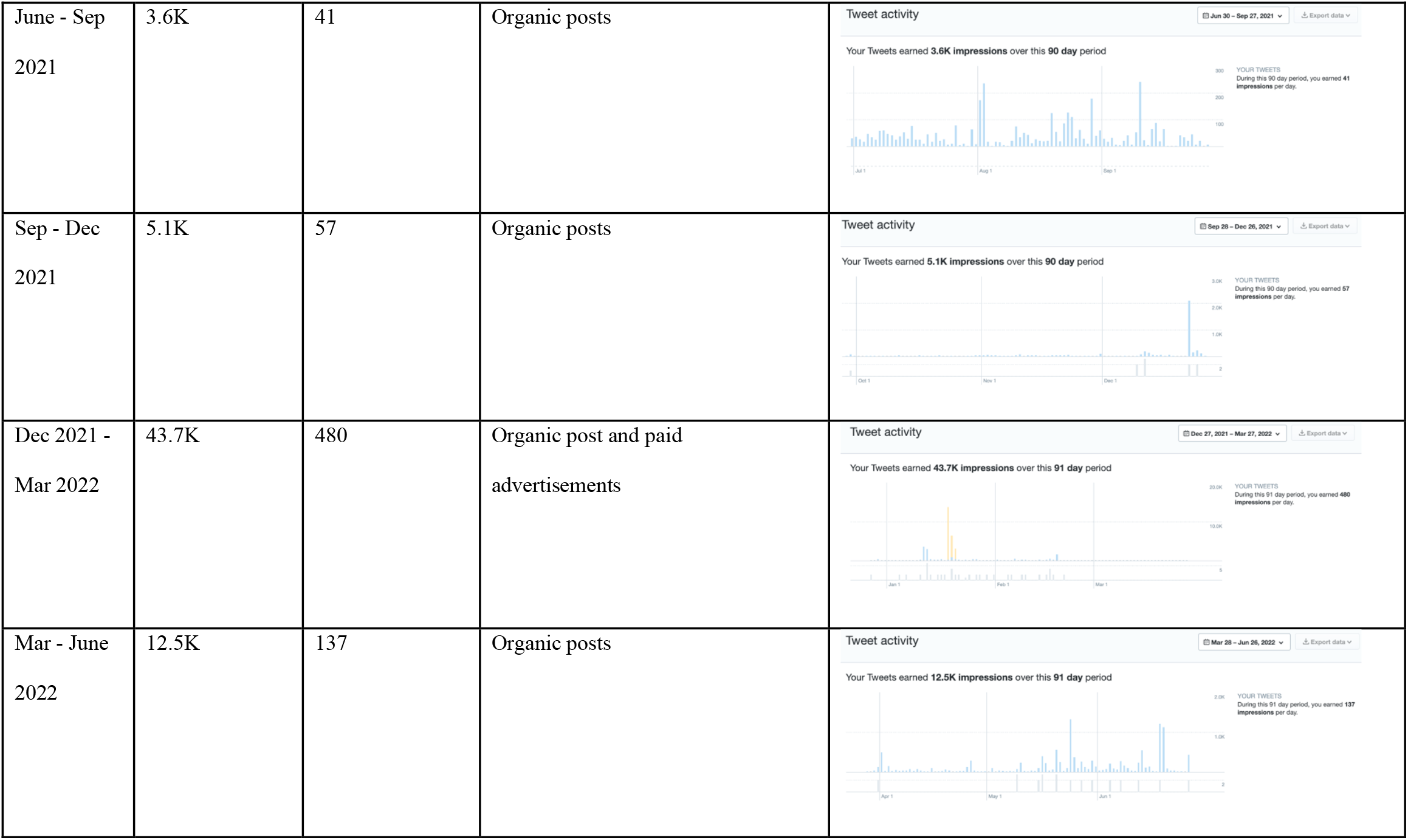
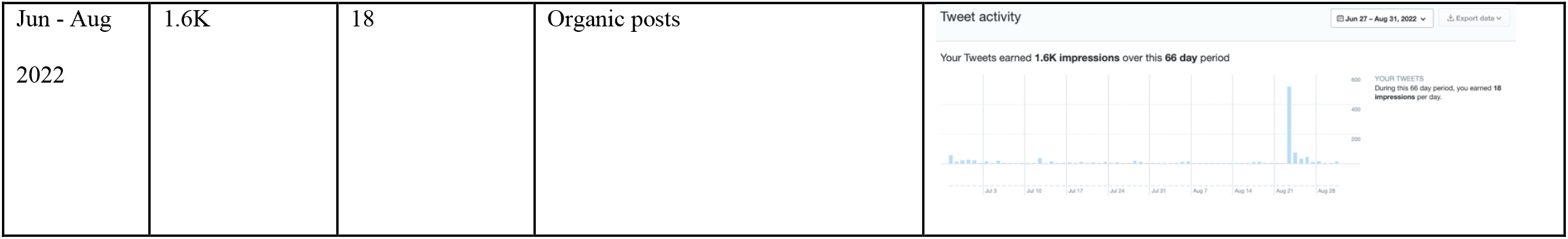
Twitter analytics over 90-day time period

### Content and impact of individual social media posts

Throughout the PSP, the top reaching post was a French-language advertisement (Figure 8). This post explicitly asked for knowledge user participation in priority-setting, included related hashtags, and had an embedded link to the French language research prioritization. This advertised post reached 13,393 users, resulting in 200 clicks on the survey link, with approximately a cost of 0.69 Canadian dollars per click. The top “organic” post was an English-language Facebook post that reached 5,415 users, and resulted in 47 link clicks. Organic posts are defined as posts for which did not have paid advertisements applied. From the top 10 posts across both Facebook and Instagram, the majority of the posts were video-based (n=9, 90%) as opposed to picture-based. Table 3 provides an overview of the top five posts, from both Facebook and Instagram, including both paid and organic posts.

**Table 3.**
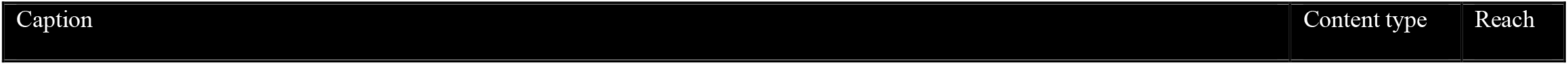
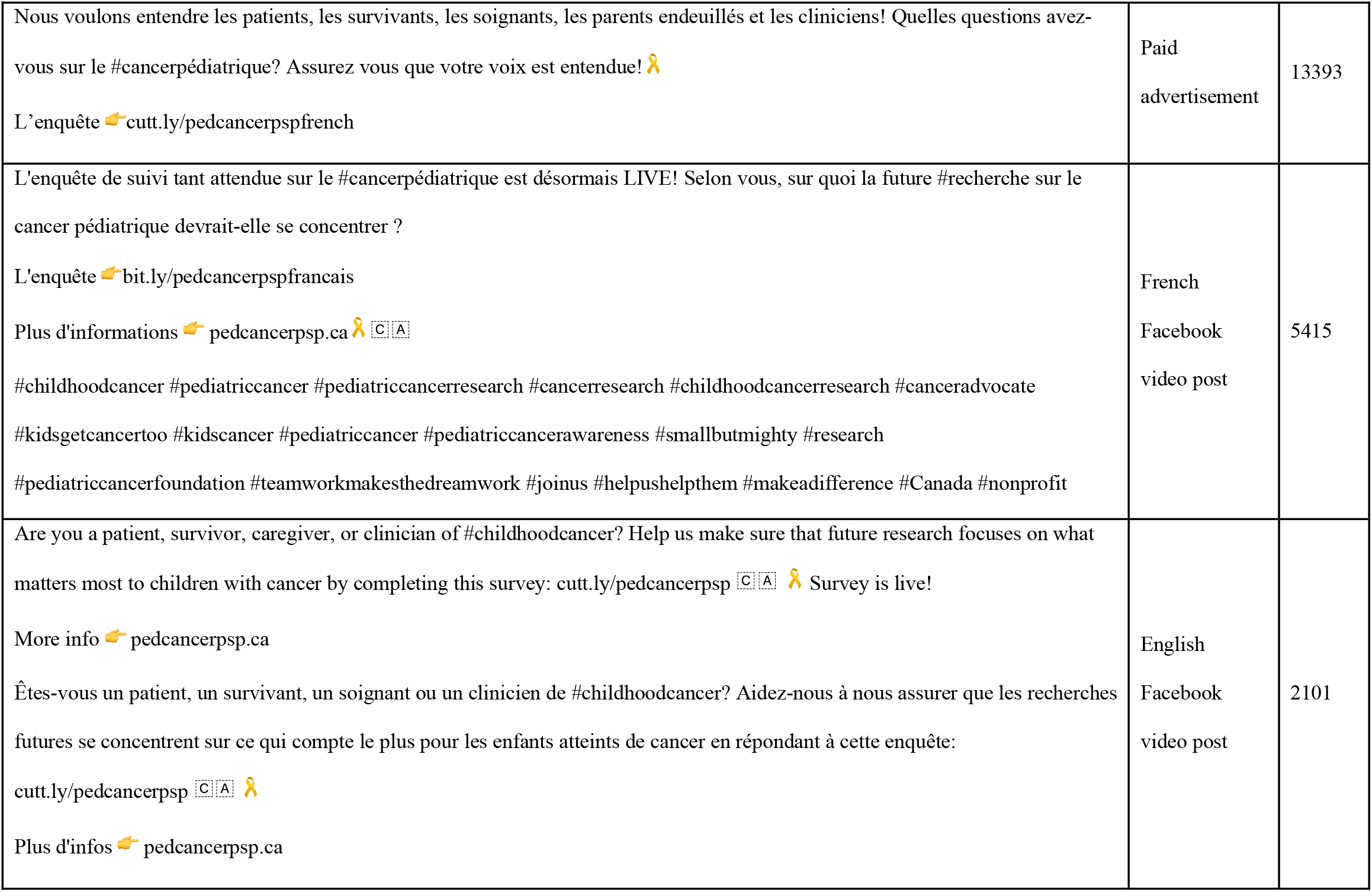
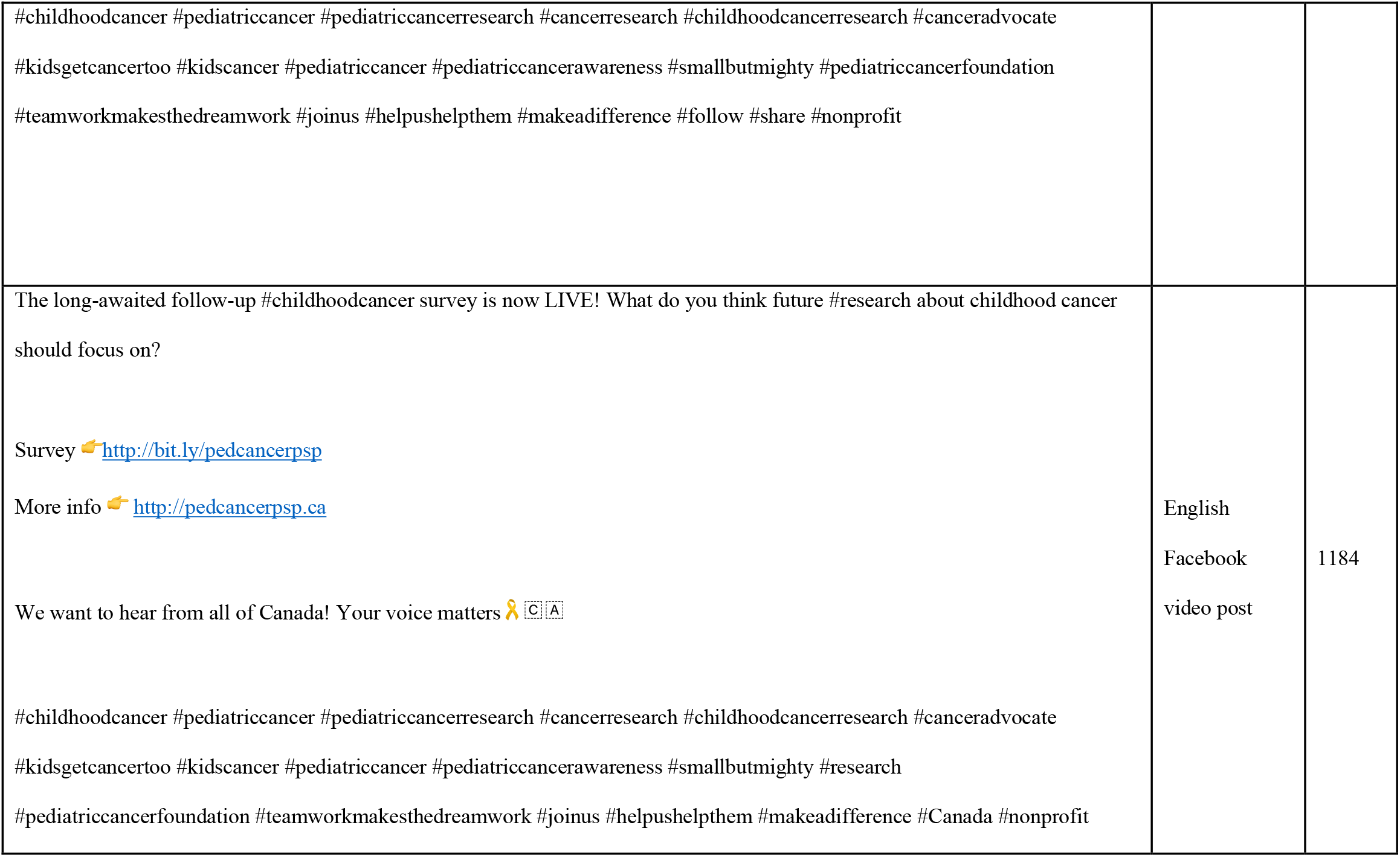
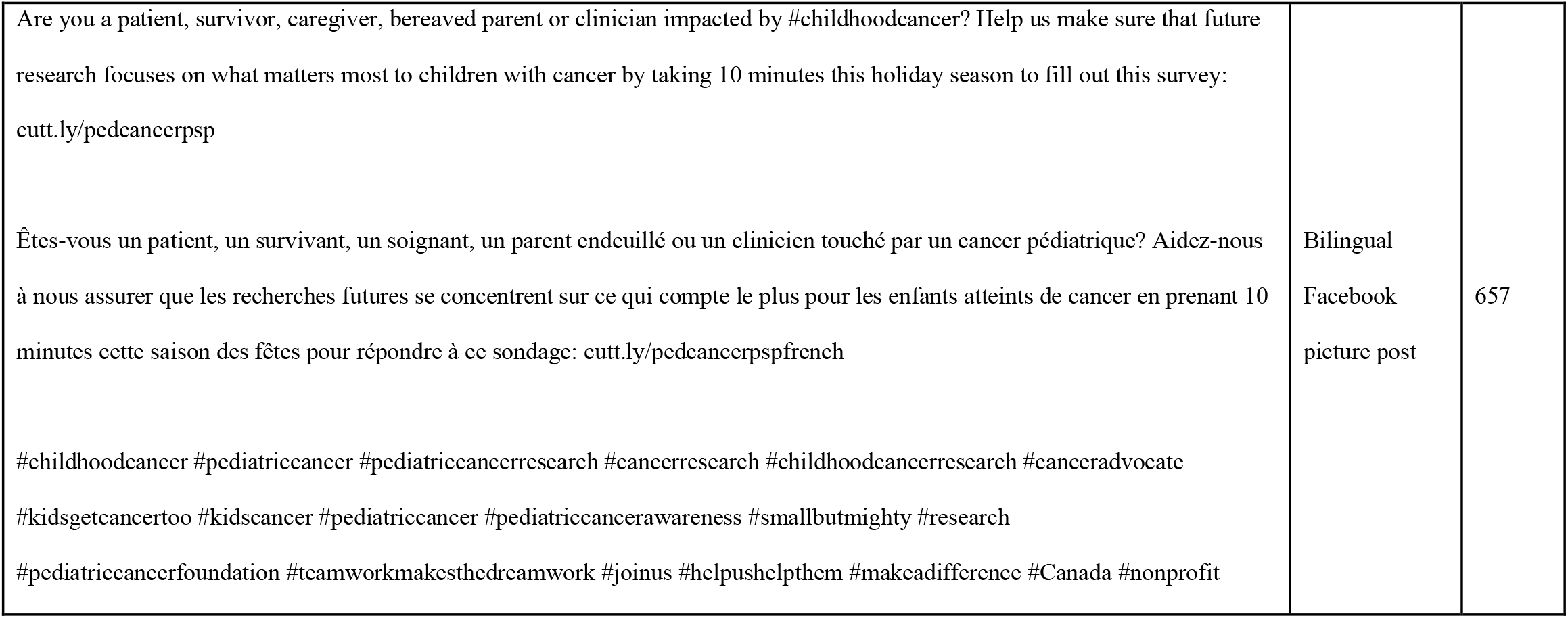
Top five posts from Facebook and Instagram

| Caption | Content type | Reach |
| --- | --- | --- |
| Nous voulons entendre les patients, les survivants, les soignants, les parents endeuillés et les cliniciens! Quelles questions avez-vous sur le #cancerpédiatrique? Assurez vous que votre voix est entendue! 🧡<br>L'enquête 📄 cutt.ly/pedcancerpspfrench | Paid advertisement | 13393 |
| L'enquête de suivi tant attendue sur le #cancerpédiatrique est désormais LIVE! Selon vous, sur quoi la future #recherche sur le cancer pédiatrique devrait-elle se concentrer ?<br>L'enquête 📄 bit.ly/pedcancerpspfrancais<br>Plus d'informations 📄 pedcancerpsp.ca 🧡 [C][A]<br>#childhoodcancer #pediatriccancer #pediatriccancerresearch #cancerresearch #childhoodcancerresearch #canceradvocate<br>#kidsgetcancertoo #kidscancer #pediatriccancer #pediatriccancerawareness #smallbutmighty #research<br>#pediatriccancerfoundation #teamworkmakesthedreamwork #joinus #helpushelpthem #makeadifference #Canada #nonprofit | French<br>Facebook<br>video post | 5415 |
| Are you a patient, survivor, caregiver, or clinician of #childhoodcancer? Help us make sure that future research focuses on what matters most to children with cancer by completing this survey: cutt.ly/pedcancerpsp [C][A] 🧡 Survey is live!<br>More info 📄 pedcancerpsp.ca<br>Êtes-vous un patient, un survivant, un soignant ou un clinicien de #childhoodcancer? Aidez-nous à nous assurer que les recherches futures se concentrent sur ce qui compte le plus pour les enfants atteints de cancer en répondant à cette enquête:<br>cutt.ly/pedcancerpsp [C][A] 🧡<br>Plus d'infos 📄 pedcancerpsp.ca | English<br>Facebook<br>video post | 2101 |
| #childhoodcancer #pediatriccancer #pediatriccancerresearch #cancerresearch #childhoodcancerresearch #canceradvocate<br>#kidsgetcancertoo #kidscancer #pediatriccancer #pediatriccancerawareness #smallbutmighty #pediatriccancerfoundation<br>#teamworkmakesthedreamwork #joinus #helpushelpthem #makeadifference #follow #share #nonprofit |  |  |
| The long-awaited follow-up #childhoodcancer survey is now LIVE! What do you think future #research about childhood cancer should focus on?<br>Survey 🖱️ http://bit.ly/pedcancerpsp<br>More info 🖱️ http://pedcancerpsp.ca<br>We want to hear from all of Canada! Your voice matters 🧡<br>#childhoodcancer #pediatriccancer #pediatriccancerresearch #cancerresearch #childhoodcancerresearch #canceradvocate<br>#kidsgetcancertoo #kidscancer #pediatriccancer #pediatriccancerawareness #smallbutmighty #research<br>#pediatriccancerfoundation #teamworkmakesthedreamwork #joinus #helpushelpthem #makeadifference #Canada #nonprofit | English<br>Facebook<br>video post | 1184 |
| Are you a patient, survivor, caregiver, bereaved parent or clinician impacted by #childhoodcancer? Help us make sure that future research focuses on what matters most to children with cancer by taking 10 minutes this holiday season to fill out this survey: cutt.ly/pedcancerpsp<br>Êtes-vous un patient, un survivant, un soignant, un parent endeuillé ou un clinicien touché par un cancer pédiatrique? Aidez-nous à nous assurer que les recherches futures se concentrent sur ce qui compte le plus pour les enfants atteints de cancer en prenant 10 minutes cette saison des fêtes pour répondre à ce sondage: cutt.ly/pedcancerpspfrench<br>#childhoodcancer #pediatriccancer #pediatriccancerresearch #cancerresearch #childhoodcancerresearch #canceradvocate #kidsgetcancertoo #kidscancer #pediatriccancer #pediatriccancerawareness #smallbutmighty #research #pediatriccancerfoundation #teamworkmakesthedreamwork #joinus #helpushelpthem #makeadifference #Canada #nonprofit | Bilingual<br>Facebook<br>picture post | 657 |

**Figure 8.**
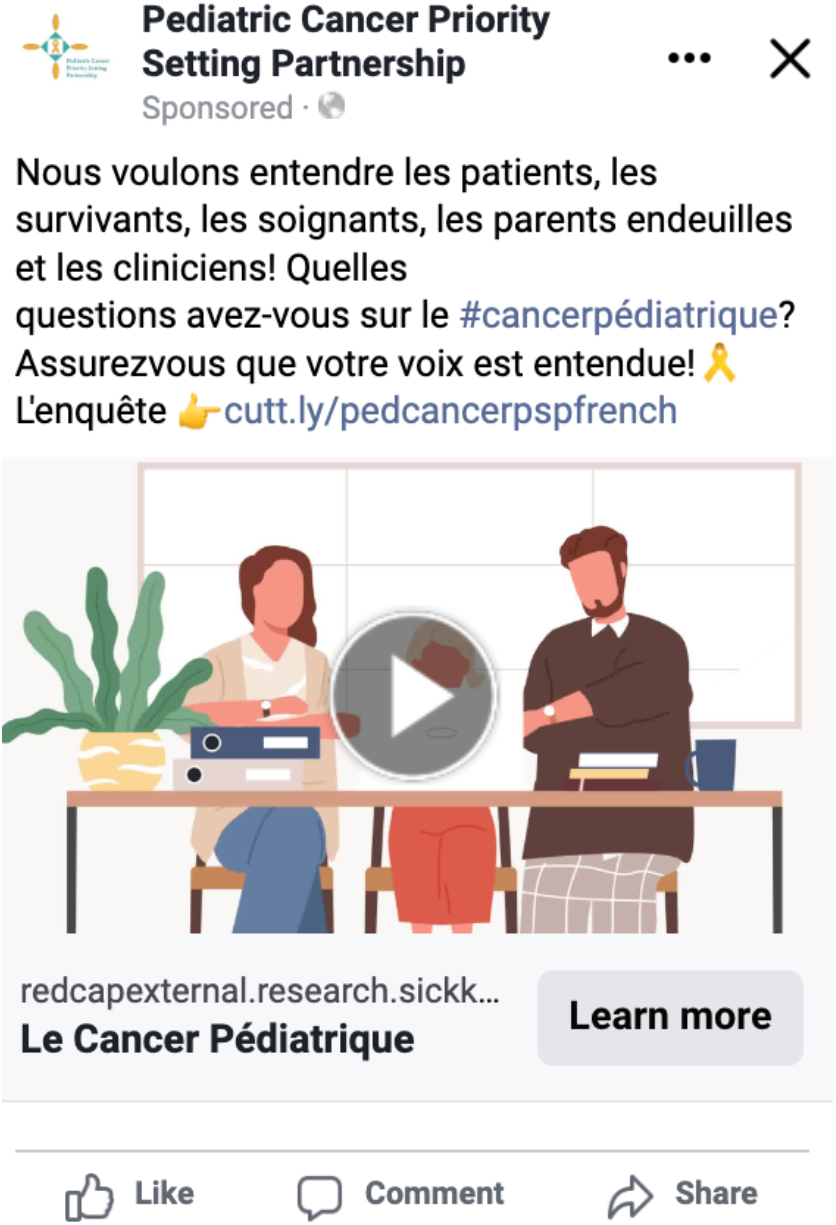
Example of French-language graphic from social media campaign

## DISCUSSION

### Principal Findings

Social media represents a promising set of tools to support health research, including the engagement of knowledge users in research priority-setting. Our research team successfully used a multi-pronged strategy leveraging social media and other online modalities to engage individuals with lived experience with childhood cancer in our pan-Canadian pediatric oncology PSP. Overall, our results demonstrate the strong utility of social media as a means to engage widespread and diverse knowledge users in PSP.

Our study website amassed 1,029 site visits, most of whom were directed to the page from our social media platforms (n=148, 32.7%). With website visitors originating from various parts of Canada (n=622, 78.8%) and the United States (n=73, 9.3%), social media campaigning has allowed us to achieve a Pan-Canadian survey by eliminating the traditional geographic barriers associated with such projects. Our campaigns on Facebook and Instagram had substantial success in reaching knowledge users, 28,641 and 2,954 people respectively across both surveys. In a similarly Pan-Canadian PSP conducted by Brockway et al., most participants were recruited through social media campaigning (n=337, 70.2%), reflecting similar effectiveness as our initial survey dissemination, which collected responses from 330 unique respondents (12). While our PSP survey recruitment was strictly limited to social media- and electronic network-based promotions, Brockway et al. integrated an in-person component where participants were informed of the study then provided with a device to record their responses (12). Though this in-person recruitment approach was inaccessible for our PSP due to the restrictions posed by the ongoing COVID-19 pandemic, the limited number of survey respondents recruited by Brockway et al. through in-person sessions (n=143; 29.8%), minimizes the potential of sampling bias in our study (12).

Furthermore, targeted paid advertising on social media platforms proved effective in increasing the reach of our social media campaign. Our posts which received a paid boost reached more users than organic posts. In fact, we received the greatest number of Facebook and Instagram impressions when paid advertisements were implemented and each paid advertisement period was accompanied by a peak in the number of survey responses collected by our team. The apparent success in terms of knowledge user engagement resulting from the use of paid advertisements is in agreement with previous research (13,14,15). Several studies demonstrate that this promotional effort results in reaching significantly more users (13,14,16). Based on our social media engagement analytics, video posts tended to reach a broader audience than image-based posts (17). Given this insight, our paid advertisements were mainly used for our video contents to optimize its effectiveness.

In addition to paid advertisements, we created social media content specifically targeting under-represented groups to encourage their participation. Throughout the social media campaign, the research team monitored social media analytics and the diversity of survey responses to identify gaps in the social media campaign and subsequently inform the content of future posts. When the research team identified lower survey response rates from people identifying with certain demographic characteristics, targeted posts calling for the involvement of these populations were made. These efforts appeared successful and correlated with an increased number of survey submissions from these groups. Additionally, to encourage pan-Canadian participation and representation from Francophone Canadians, our entire social media campaign was bilingual with both French and English language posts made. In these ways, we employed a tailored approach to reach specific populations on social media and avoided utilizing a “one-size-fits-all” dissemination technique. In similar studies assessing the application of social media in health research, tailoring social media content to specific populations have shown to enhance engagement and users’ experience (18). Reflecting the findings of our study, targeting content to particular groups has also been identified as an effective means to fill gaps in the reach of priority setting partnership survey promotions (19,20).

We also collaborated with relevant organizations to promote our social media campaign. This strategy is embedded in the James Lind Alliance methodology and was a key factor for success in research priority-setting as identified in our previous scoping review (7). In health research, there are challenges associated with approaching members of the general public due to the inherent power dynamic that exists with healthcare providers (21). Thus, partnering with external organizations and communities that are known and trusted by a social media target audience can help to build campaign credibility, overcome historical challenges to research participation (22), and thereby increase the number of survey responses. Although, in principle, the possibility of content spreading widely and organically via social media exists, this is difficult without an established and well-connected social media presence (16). Given that our project involved building and utilizing new social media channels with no prior following, it was particularly important to leverage relationships with external organizations and communities to promote our research prioritization surveys via their established networks. Our findings show that involving external individuals and organizations in promotion directly translated to a surge in the survey response rate during the course of both the initial and interim surveys. External organizations were more inclined to help with promotion when provided with a template package, including tailored graphics, pre-written messaging, sample newsletter graphics, and draft emails. Contacting organizational partners before the study start date, sending regular, brief reminders throughout the study period, and providing some degree of mutual benefit (e.g., study progress updates, providing a summary of study results) to encourage their support have proven beneficial in leveraging partnerships and furthering our outreach. Table 4 provides an overview of the specific recommendations derived from our social media campaign.

**Table 4.**
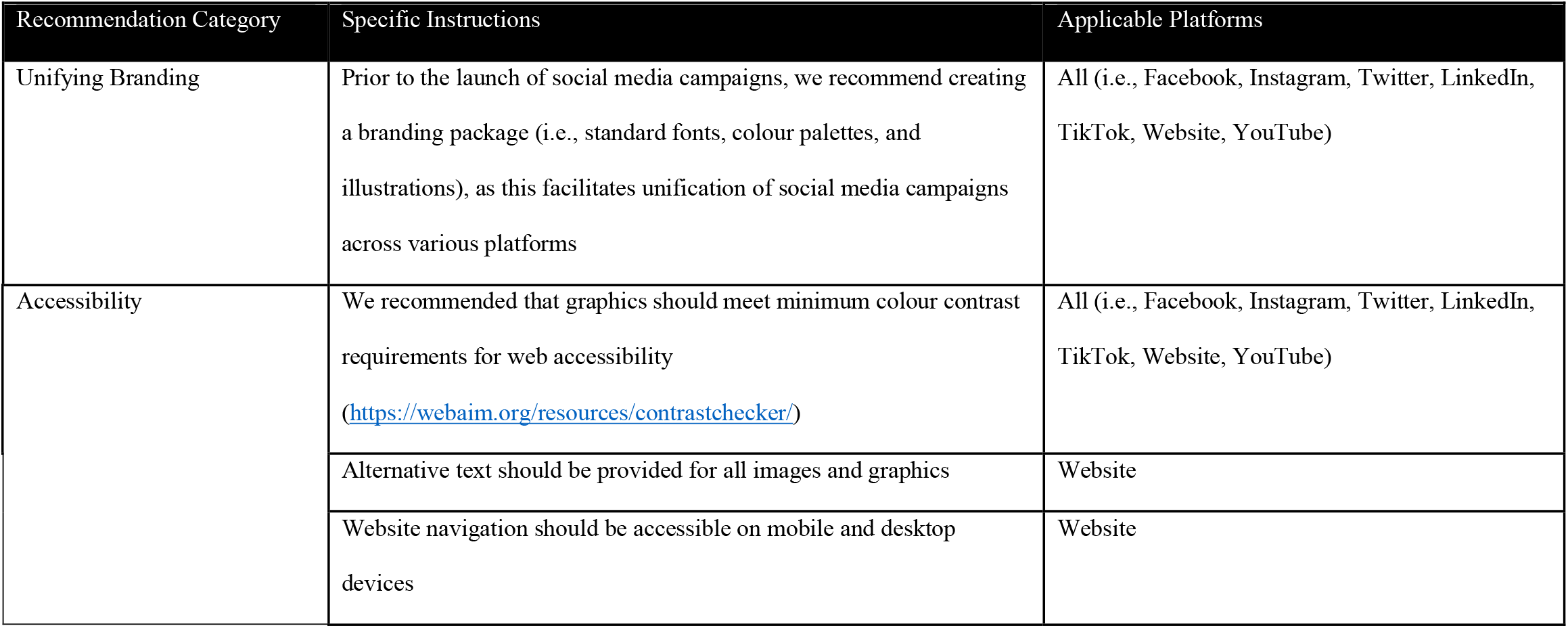
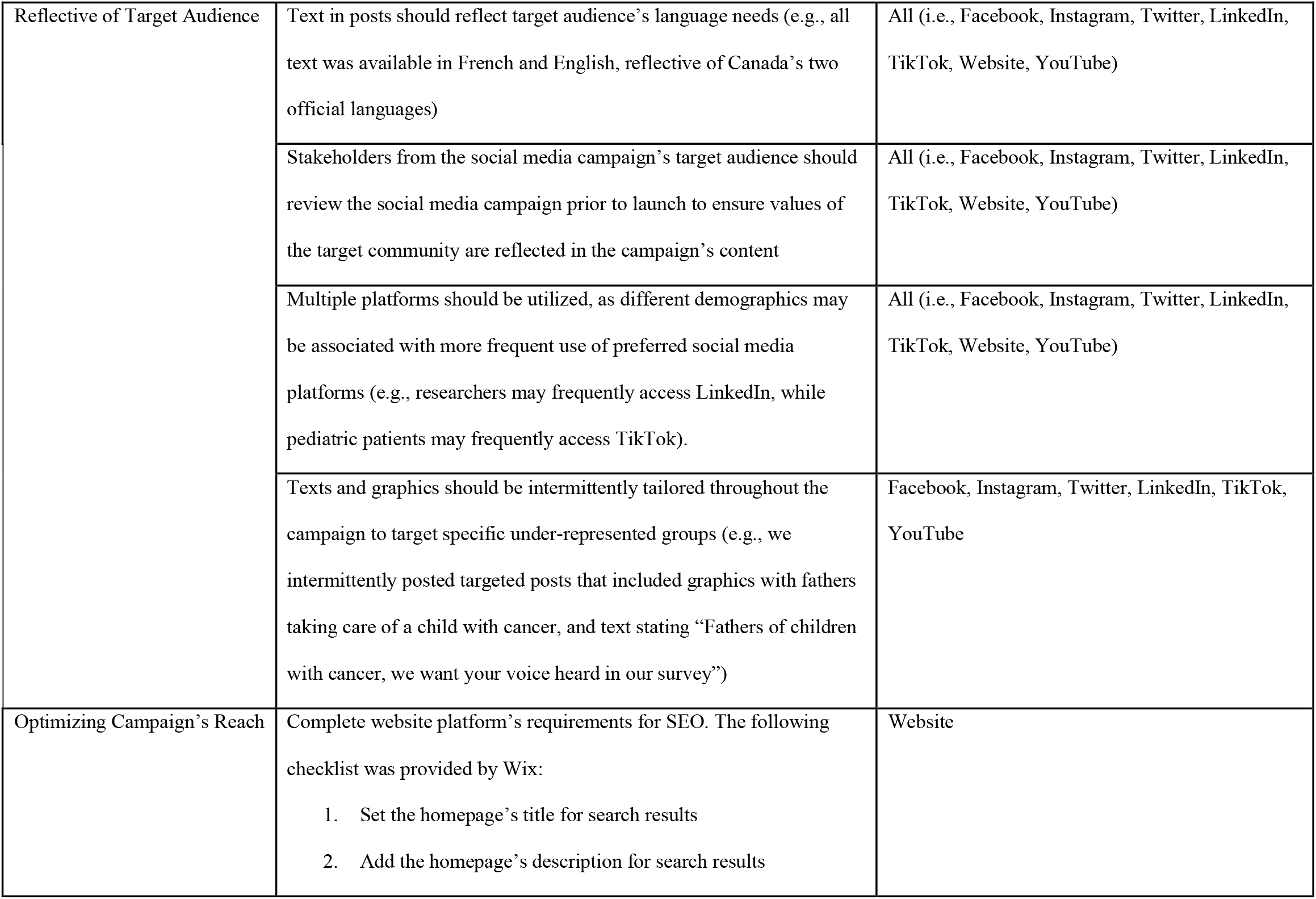
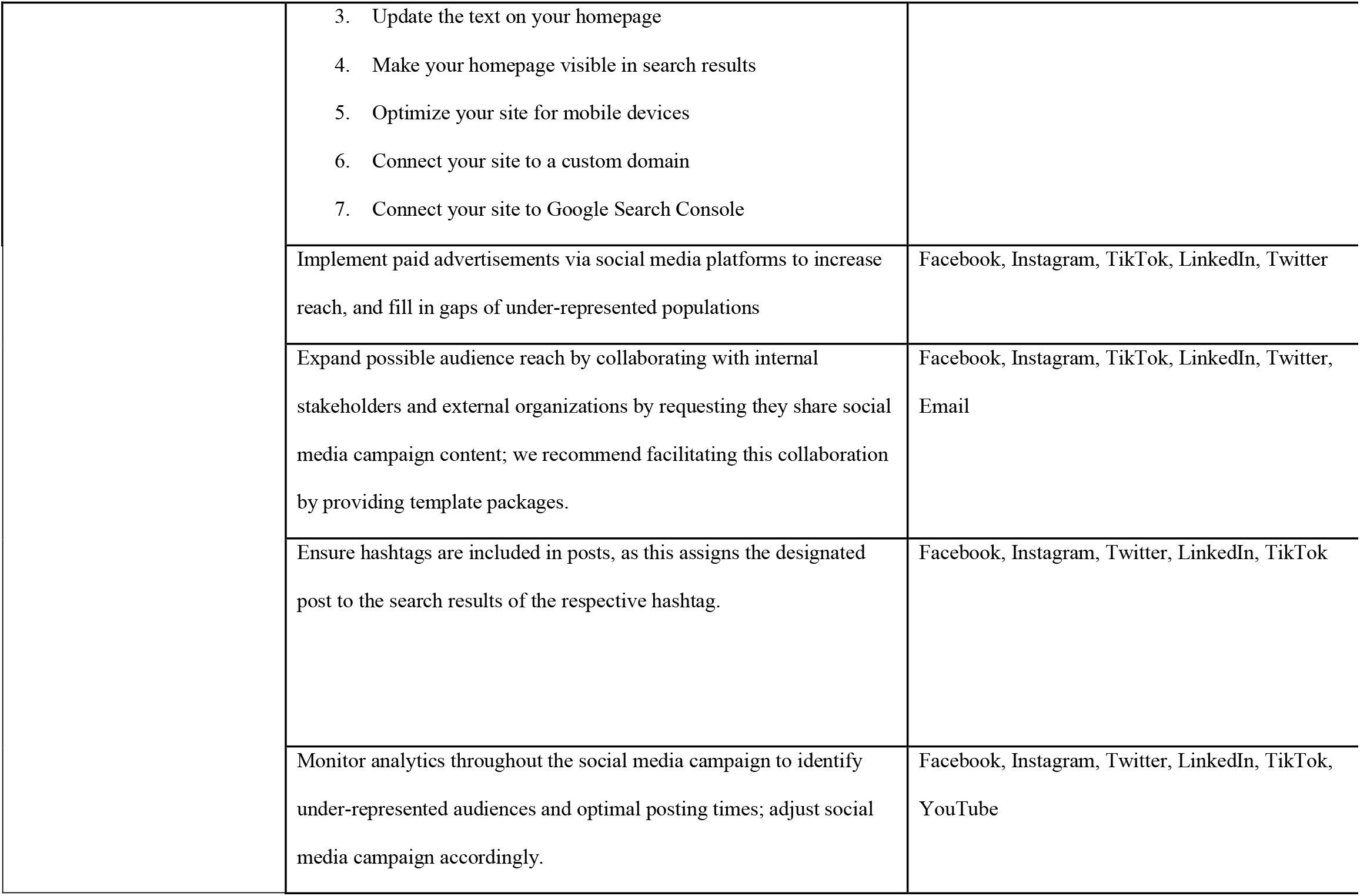
Recommendations for future research-based social media campaign

| Recommendation Category | Specific Instructions | Applicable Platforms |
| --- | --- | --- |
| Unifying Branding | Prior to the launch of social media campaigns, we recommend creating a branding package (i.e., standard fonts, colour palettes, and illustrations), as this facilitates unification of social media campaigns across various platforms | All (i.e., Facebook, Instagram, Twitter, LinkedIn, TikTok, Website, YouTube) |
| Accessibility | We recommended that graphics should meet minimum colour contrast requirements for web accessibility<br>(https://webaim.org/resources/contrastchecker/) | All (i.e., Facebook, Instagram, Twitter, LinkedIn, TikTok, Website, YouTube) |
|  | Alternative text should be provided for all images and graphics | Website |
|  | Website navigation should be accessible on mobile and desktop devices | Website |
| Reflective of Target Audience | Text in posts should reflect target audience's language needs (e.g., all text was available in French and English, reflective of Canada's two official languages) | All (i.e., Facebook, Instagram, Twitter, LinkedIn, TikTok, Website, YouTube) |
|  | Stakeholders from the social media campaign's target audience should review the social media campaign prior to launch to ensure values of the target community are reflected in the campaign's content | All (i.e., Facebook, Instagram, Twitter, LinkedIn, TikTok, Website, YouTube) |
|  | Multiple platforms should be utilized, as different demographics may be associated with more frequent use of preferred social media platforms (e.g., researchers may frequently access LinkedIn, while pediatric patients may frequently access TikTok). | All (i.e., Facebook, Instagram, Twitter, LinkedIn, TikTok, Website, YouTube) |
|  | Texts and graphics should be intermittently tailored throughout the campaign to target specific under-represented groups (e.g., we intermittently posted targeted posts that included graphics with fathers taking care of a child with cancer, and text stating "Fathers of children with cancer, we want your voice heard in our survey") | Facebook, Instagram, Twitter, LinkedIn, TikTok, YouTube |
| Optimizing Campaign's Reach | Complete website platform's requirements for SEO. The following checklist was provided by Wix:<br>1. Set the homepage's title for search results<br>2. Add the homepage's description for search results | Website |
|  | 3. Update the text on your homepage<br>4. Make your homepage visible in search results<br>5. Optimize your site for mobile devices<br>6. Connect your site to a custom domain<br>7. Connect your site to Google Search Console |  |
|  | Implement paid advertisements via social media platforms to increase reach, and fill in gaps of under-represented populations | Facebook, Instagram, TikTok, LinkedIn, Twitter |
|  | Expand possible audience reach by collaborating with internal stakeholders and external organizations by requesting they share social media campaign content; we recommend facilitating this collaboration by providing template packages. | Facebook, Instagram, TikTok, LinkedIn, Twitter, Email |
|  | Ensure hashtags are included in posts, as this assigns the designated post to the search results of the respective hashtag. | Facebook, Instagram, Twitter, LinkedIn, TikTok |
|  | Monitor analytics throughout the social media campaign to identify under-represented audiences and optimal posting times; adjust social media campaign accordingly. | Facebook, Instagram, Twitter, LinkedIn, TikTok, YouTube |

### Limitations

There were limitations of this study and the corresponding social media campaign. We observed a substantial gap between the number of visitors to our social media platforms and those that proceeded to open and complete the survey. This is a commonly cited limitation in PSP, where the social media campaign reach is disproportionately greater than the survey response rates (16). Although the platform analytics (i.e., number of follows, comments, and likes on posts) were often used as an indication of our survey engagement, these social media insights were not representative of the sample that proceeded to open the survey link and complete the survey. While clear instructions on accessing and completing the survey were provided in each social media post, potential contributors to the low response rate may have stemmed from uncertainty about eligibility or possible difficulties faced within the actual survey. Elaborating on these details and improving within survey navigation may have yielded greater response rates from those interacting with our social media posts.

Another limitation of social media usage in priority-setting research includes the uncertainty of who is being captured through the posts (23). Social media–based methods may unexpectedly include or exclude the research priority perspectives of certain groups (7). For our survey, the team ensured that social media posts were accompanied with a clear definition of the survey respondent eligibility criteria. Additionally, the survey was prefaced by a screening question intended to filter out ineligible respondents. Despite these efforts, we recognize that the widespread reach of social media contents inherently limits the control we have over the survey respondents. Individuals that do not fit our PSP eligibility criteria were still able to complete the survey on their own accord. As such, social media-based recruitment for PSPs constraints researchers’ abilities to appropriately screen survey respondents. This is largely due to the limited control over who sees the social media content once posted (24).

Despite our attempts to achieve a pan-Canadian survey, we experienced difficulty engaging certain geographic regions. Particularly, it was challenging during the initial survey term to reach Francophones situated in Quebec. While we employed targeted posts and paid advertisements towards this population, these efforts proved futile, as evidenced by the low respondent rate from Quebec residents in the initial survey (n=24, 7.3%). Similarly, across both surveys, there were significantly more female respondents than male (n=441 and n=82, respectively). Thus, to promote a more diverse respondent demographic, we connected with external groups (i.e., Quebec-based cancer organizations and private Facebook parent groups). This reinforces the importance of supplementing social media posts with additional engagement strategies, including actively connecting with external stakeholders to promote the content.

Although social media campaigning efforts were prioritized on Facebook, Twitter, Instagram, and the study website, we expanded our outreach to YouTube and TikTok—some of the most utilized social media platforms among young people (25,26). Given that our target audience included patients of childhood cancer, we believed that availing to platforms that are more popular among the younger demographic (i.e., 0-18 years of age) would help better engage this population.

Despite our efforts to attract the younger age groups, we had limited data on the impact of these strategies. Specifically, given the short duration in which our YouTube and TikTok accounts were active, there was limited insight collected on the effectiveness of these platforms in reaching younger aged survey respondents. Recognizing that social media preferences among young people are constantly evolving, a continued understanding of forthcoming platforms among the younger population is warranted (27).

### Future Research

We recognize that little is known on the operation of social media algorithms. Further research is needed to understand how social media algorithms can influence the capacity of social media recruitment to capture representative samples. More research is also needed to understand which social media strategies and dissemination techniques are likely to be successful for research prioritization efforts, with the understanding that these strategies and techniques are likely to change over time as new social media platforms and features become available.

Given the relatively nuanced role of social media in research methods, navigating the new territory for ethical approval warrants further research. Many of the ethics requirements of customary recruitment techniques are difficult to translate to research using social media given the vast potential reach (16). A particular ethical concern posed by social media recruitment relates to privacy. Privacy risk has been cited as a primary concern among research participants, particularly for the general public with limited research background (28,29). In PSP exercises, the engagement of patients and their families are of utmost importance. Thus, gaining exposure and continuing to evaluate aspects of social media from the context of privacy and users’ comfort levels is critical to inform the development of efficient and effective strategies to connect with the general public. In doing so, social media-based research methodologies can be placed in proper ethical perspective and be utilized as a valuable recruitment tool.

## CONCLUSION

The engagement of people with lived healthcare experiences in research priority-setting is critically important. However, the best methods for engaging knowledge users are scantly identified in literature. Our study evaluated social media as a novel approach to engage pediatric cancer patients, survivors, their family caregivers, and healthcare providers in the development of a Canadian research agenda. Diversifying recruitment across multiple platforms increased response rates and improved the reach in an efficient manner. Utilizing paid advertisements, tailoring social media content to specific populations, and circulating promotional material to partner organizations have proven effective in increasing engagement in our PSP. Further investigation of social media algorithms and dissemination techniques is needed to understand how to increase representation among survey respondents. Consideration of the privacy implications of social media use for research is also needed. Ultimately, continued evaluation of novel tools to enable inclusive priority-setting is needed to amplify the voices of those with lived experience as it pertains to the next scientific efforts.

## Data Availability

All relevant data are within the manuscript and its Supporting Information files.

## DECLARATIONS

Ethics approval for this study was received from the SickKids Research Ethics Board (REB). Consent for publication was not applicable. All data generated or analysed during this study are included in this published article. The authors declare that they have no competing interests. This study was funded by the Canadian Institute of Health and Research (CIHR). SS created graphics and the branding package, led the launch of the social media campaign and conducted data analyses. RA created and posted graphics on social media. KH monitored survey respondents’ demographics. EH and LJ headed the PSP and worked with the steering group and authors of this paper to build a productive social media strategy to meet the larger PSP’s goals. All authors read and approved the final manuscript.

## ABBREVIATIONS

BIPOC: Black, Indigenous, People of Colour
CIHR: Canadian Institute of Health and Research
COVID-19: Coronavirus disease 2019
GIFs: Graphic interchange format
PSP: Priority setting partnership
REB: Research Ethics Board
SEO: Search optimization checklist

